# Insight into risk associated phenotypes behind COVID-19 from phenotype genome-wide association studies

**DOI:** 10.1101/2023.05.09.23289706

**Authors:** Zhongzhong Chen, Koichi Matsuda

## Abstract

Long COVID presents a complex and multi-systemic disease that poses a significant global public health challenge. Symptoms can vary widely, ranging from asymptomatic to severe, making the condition challenging to diagnose and manage effectively. Furthermore, identifying appropriate phenotypes in genome-wide association studies of COVID-19 remains unresolved. This study aimed to address these challenges by analyzing 220 deep-phenotype genome-wide association data sets (159 diseases, 38 biomarkers and 23 medication usage) from BioBank Japan (BBJ) (n=179,000), UK Biobank and FinnGen (n=628,000) to investigate pleiotropic effects of known COVID-19 risk associated single nucleotide variants. Our findings reveal 32 different phenotypes that share the common genetic risk factors with COVID-19 (*p* < 7.6×10^−11^), including two diseases (myocardial infarction and type 2 diabetes), 26 biomarkers with seven categories (blood cell, metabolic, liver-related, kidney-related, protein, inflammatory and anthropometric), and four medications (antithrombotic agents, HMG CoA reductase inhibitors, thyroid preparations and anilides). As long COVID continues to coexist with humans, our results highlight the need for targeted screening to support specific vulnerable populations to improve disease prevention and healthcare delivery.

---

Long COVID, also known as “post-acute sequelae of COVID-19”, has been identified as the next public health concern. This debilitating illness is estimated to affect more than 200 symptoms (*1*), and may occur in about 12.5% (1/8) people with COVID-19 in the general population (*2*). The current diagnostic and treatment options are inadequate, and the underlying biological mechanisms need to be further addressed (*1*). Through genome-wide association studies (GWASs), more than 40 single nucleotide variants (SNPs) involving 35 genes have been identified as potential contributors to susceptibility to SARS-CoV-2 infection or risk of severe COVID-19 disease (*3*). While translational studies of GWAS findings may provide novel insights into pathophysiology, there remains limited understanding of the pleiotropic effects between genetic predisposition, complications, biomarkers and medications for guiding long-term follow-up and life care of COVID-19 patients.

Phenome-wide association studies (PheWASs) based on population have opened up new avenues for formulating effective biomedical hypotheses. In this study, we delved into the pleiotropic loci of known single nucleotide variants (SNPs) (*3*) that represent either susceptibility to SARS-CoV-2 infection or risk of severe COVID-19 disease. To do this, we analyzed 220 deep-phenotype genome-wide association data sets with deep-phenotype information, comprising 159 diseases, 38 biomarkers and 23 medication usage from BioBank Japan (BBJ) (n=179,000), UK Biobank and FinnGen (n_total_=628,000) (*4*).

First, using a comprehensive PheWAS analysis of the genome-wide landscape, we have identified two genetic associations with myocardial infarction and type 2 diabetes that indicate a potential link to COVID-19 infection. Our diagnosis code-focused PheWAS (Figure 1) revealed a significant association between rs912805253 at the *OAS1* locus and myocardial infarction (MI) (ICD-10 code I21/I22/I23/I24/I25) in both the BBJ and European populations [BBJ population: odds ratio (OR) = 1.09, 95% confidence interval (CI) = 1.06-1.12, *p* = 3.3×10^−11^; European population: OR = 1.04, 95% CI = 1.02-1.07, *p* = 3.4×10^−4^].

**Figure 1.**
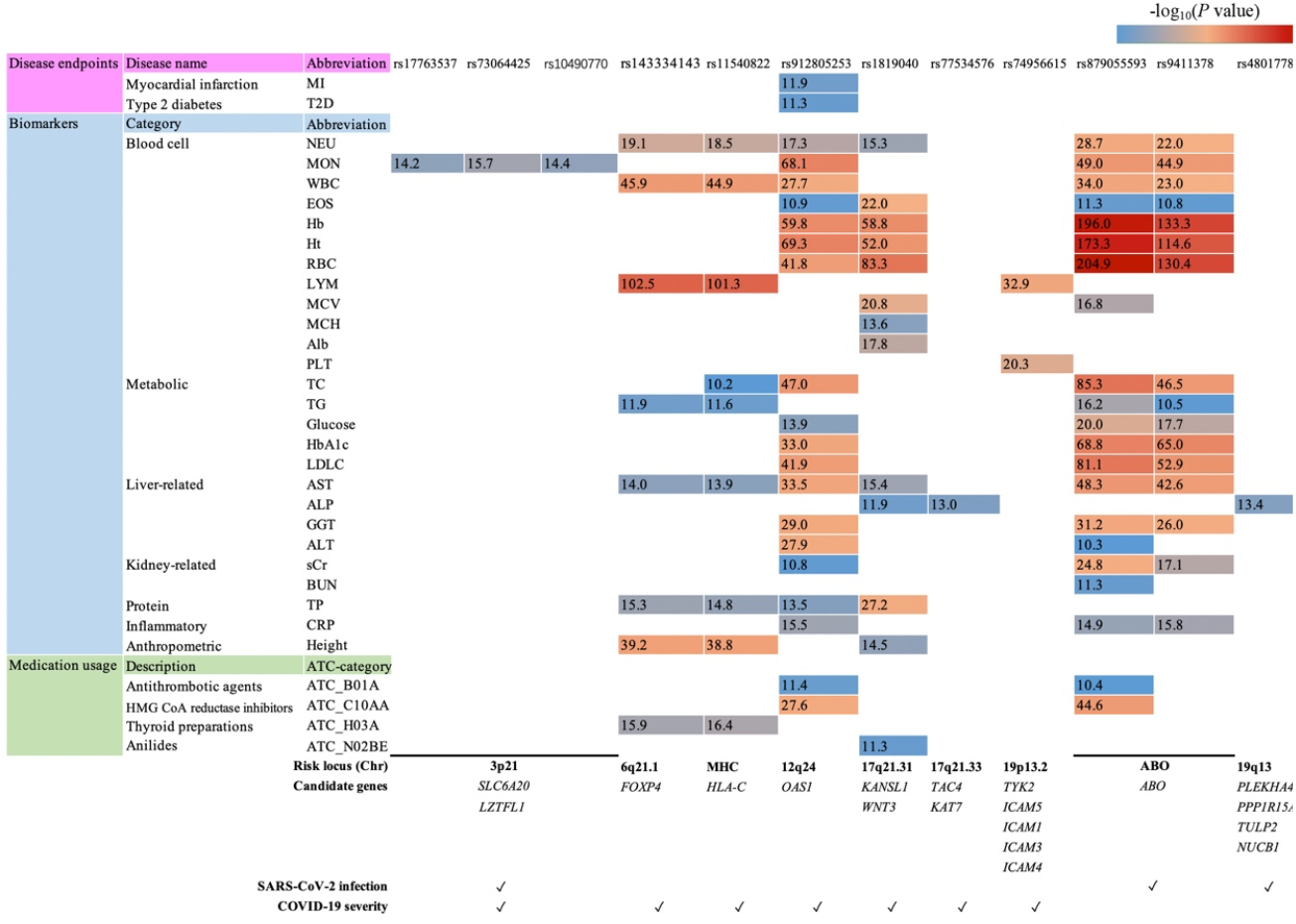
The landscape of associations with diseases, clinical measures (biomarkers) and medication usage for genetic risk loci from COVID-19 GWASs. The association results only considered the SNPs reached the Bonferroni-corrected threshold of <7.6×10^−11^, using -log10(p value). Risk locus, candidate genes and their representation for SARS-CoV-2 infection and severe COVID-19 with complications from Karlsen’s summary (*3*). Abbreviations and full names for diseases and biomarkers: MI, myocardial infarction; T2D, type 2 diabetes; NEU, neutrophil; MON, monocyte; WBC, white blood cell; EOS, eosinophil; Hb, hemoglobin; Ht, hematocrit; RBC, red blood cell; LYM, lymphocyte; MCV, mean corpuscular volume; MCH, mean corpuscular hemoglobin; Alb, albumin; PLT, platelet; TC, total cholesterol; TG, triglycerides; LDLC, LDL-cholesterol; AST, aspartate transaminase; ALP, alkaline phosphatase; GGT, γ-glutamyl transpeptidase; ALT, alanine aminotransferase; sCr, serum creatinine; BUN, blood urea nitrogen; TP, total protein; CRP, C-reactive protein.

When combining subjects both from BBJ and European population, the OR for rs912805253 with MI was 1.07 (95% CI = 1.05-1.08, *p* = 1.3×10^−12^). Furthermore, rs912805253 was also significantly associated with type 2 diabetes (T2D) (ICD-10 code E11) in both populations [BBJ population: odds ratio (OR) = 1.04, 95% confidence interval (CI) = 1.02-1.06, *p* = 3.7×10^−5^; European population: OR = 1.05, 95% CI = 1.03-1.07, *p* = 2.4×10^−8^]. When combining subjects from both population, the OR for rs912805253 with T2D was 1.05 (95% CI = 1.03-1.06, *p* = 5.6×10^−12^). Clinical studies (cohorts) have suggested and increased risk of long-term cardiovascular outcomes (*5*) and type 2 diabetes (*6*) after COVID-19 infection, but their genetic risk factors are still unclear. Our findings shed light on the potential genetic link between COVID-19 infection and these diseases, providing a valuable contribution to this field.

Second, our findings provide compelling evidence of shared genetic risk factors across 26 clinical lab-measurement, particularly among seven blood cell biomarkers (neutrophil, monocyte, white blood cell, eosinophil, hemoglobin, hematocrit and red blood cell), two metabolic biomarkers (total cholesterol and triglycerides), one liver-related biomarker (aspartate transaminase) and one protein biomarker (total protein). Based on our analysis using the clinical lab-measurement PheWAS, we identified 86 significant associations between 12 SNPs and 26 biomarkers (7.6×10^−11^) (Figure 1). Among these biomarkers, blood cell measurements emerged as one of the top associated categories, followed by metabolic, liver-related, kidney-related, protein, inflammatory, anthropometric. Within the blood cell type category, neutrophil (NEU) (rs912805253, *p* = 4.9×10^−18^; rs879055593, *p* = 1.9×10^−29^; rs9411378, *p* = 1.1×10^−22^; rs1819040, *p* = 5.0×10^−16^; rs11540822, *p* = 3.1×10^−19^; rs143334143, *p* = 8.7×10^−20^), monocyte (MON) (rs912805253, *p* = 8.2×10^−69^; rs879055593, *p* = 9.7×10^−50^; rs9411378, *p* = 1.3×10^−45^; rs17763537, *p* = 7.0×10^−15^; rs73064425, *p* = 2.2×10^−16^; rs10490770, *p* = 3.8×10^−15^), and white blood cell (WBC) (rs912805253, *p* = 1.8×10^−28^; rs879055593, *p* = 9.4×10^−35^; rs9411378, *p* = 9.7×10^−24^; rs11540822, *p* = 1.2×10^−45^; rs143334143, *p* = 1.2×10^−46^) were associated with five or more COVID-19 risk associated SNPs. Among metabolic biomarkers, total cholesterol (TC) (rs912805253, *p* = 1.1×10^−47^; rs879055593, *p* = 4.5×10^−86^; rs9411378, *p* = 3.2×10^−47^; rs11540822, *p* = 6.2×10^−11^) and triglycerides (TG) (rs879055593, *p* = 6.9×10^−17^; rs9411378, *p* = 3.4×10^−11^; rs11540822, *p* = 2.8×10^−12^; rs143334143, *p* = 1.2×10^−12^) were each associated with four SNPs. Finally, aspartate transaminase (AST) in the kidney-related category was enriched for six COVID-19 risk associated SNPs (rs912805253, *p* = 3.3×10^−34^; rs879055593, *p* = 5.4×10^−49^; rs9411378, *p* = 2.5×10^−43^; rs1819040, *p* = 4.3×10^−16^; rs11540822, *p* = 1.3×10^−14^; rs143334143, *p* = 1.1×10^−14^). The identification of shared genetic risk factors across multiple clinical lab-measurements highlights the importance of understanding the interplay between various biological pathways and how they contribute to COVID-19 severity. By analyzing an individual’s genetic profile and clinical lab-measurements, healthcare providers may be able to better predict the patient’s risk of developing severe symptoms or long-term complications from COVID-19. These findings could help inform treatment decisions and prioritize high-risk patients for early intervention.

Third, in our PheWAS analysis of medication usage, we found that antithrombotic agents (rs912805253, *p* = 3.6×10^−12^; rs879055593, *p* = 4.1×10^−11^), HMG CoA reductase inhibitors (rs912805253, *p* = 2.8×10^−28^; rs879055593, *p* = 2.5×10^−45^), thyroid preparations (rs11540822, *p* = 3.7×10^−17^; rs143334143, *p* = 1.3×10^−16^), and anilides (rs1819040, *p* = 4.6×10^−12^) have shared genetic risk factors with COVID-19 (Figure 1). This suggests that individuals with a genetic predisposition to these medications may be more vulnerable to severe COVID-19 symptoms. This findings highlight the importance of personalized treatment and management of COVID-19 based on an individual’s genetic background. Furthermore, the discovery of common genetic risk factors between certain medications and COVID-19 opens up new possibilities for developing targeted therapies. By identifying the underlying genetic mechanisms, researchers could develop new medications that could modify these risk factors and potentially improve patient outcomes. In the future, the personalized approach to COVID-19 treatment and the development of new medications may greatly enhance our ability to combat this pandemic.

The current diagnostic and treatment protocols for long COVID are inadequate to fully comprehend its overlap with other diseases, the variable onset of symptoms, as well as the effects of vaccinations (*1*). Definitive diagnostic criteria for long COVID have yet to be established (*7*). Establishing a comprehensive surveillance system is imperative to effectively manage the ongoing presence of long COVID. Vaccination remains the ultimate protection against SARS-CoV-2 infection. To address these gaps, our study characterized the risk associated phenotypes behind COVID-19 by incorporating PheWASs data sets from BBJ and European biobank (*4*), and summarized data from published and peer-reviewed GWAS articles on COVID-19 (*3*). We identified 32 significant associated phenotypes across two diseases, 26 biomarkers, and four medications after conservative Bonferroni multiple testing correction (*p* < 7.6×10^−11^). Our findings provide valuable insights into the pathophysiology of COVID-19 infection, its complications, treatment, genetic risk prediction and lifelong care. This study could aid in the identification and development of new treatment options.

## Data Availability

All data produced in the present study are available upon reasonable request to the authors

## Acknowledgments

The authors wish to thank Dr. Aiko Kurosaka from the University of Tokyo for providing valuable insights during our analysis.

## Author contributions

Z.C. designed the study and wrote the paper; K.M. coordinated the project; all authors reviewed the paper and contributed to scientific content.

